# A multilevel modeling of factors on Sulfadoine-Pyrimethanie uptake in Cameroon: Evidence from national cross-sectional survey

**DOI:** 10.1101/2024.05.28.24308039

**Authors:** Linus Baatiema, Ambrose Naawa, Anthony M. Tampah-Naah, Jerry P. K Ninnoni, Kennedy A. Alatinga, Mukaila Mumuni Zankawah, Gladys Sufuyan, Munawar Harun Koray

## Abstract

**Introduction:** Pregnant women worldwide face significant risks from malaria, which can adversely affect both mother and baby. To combat malaria in Cameroon, the WHO, in 2004, recommended a strategy including intermittent preventive treatment during pregnancy (IPTp) using sulfadoxine-pyrimethamine (SP), insecticide-treated nets (ITNs), and proper management for clinical malaria and anaemia. This study explores the prevalence and predictors of IPTp-SP uptake across individual, community, and regional levels in Cameroon.

**Material and Method:** We utilized data from the 2018 Cameroon Demographic and Health Survey (CDHS) to analyze IPTp-SP uptake employing three-level multi-level models to capture individual, community, and regional influences. Responses from 4,823 women were analyzed, with results presented in 95% credible intervals.

**Results:** Women aged 35-39 were more likely to receive at least 3 doses of IPTp-SP during their last pregnancies compared to those aged 45-49 (aOR=1.92, 95% Crl=1.08-3.41). Women without formal education were less likely to have optimal IPTp-SP uptake (aOR=0.81, 95% Crl=0.70-0.92) compared to those with higher education. The wealthiest women and those attending at least eight ANC visits had higher odds of optimal IPTp-SP uptake (aOR=1.47, 95% Crl=1.20-2.09 and aOR=1.97, 95% Crl=1.25-3.12, respectively). Additionally, those with health insurance, urban residents, and moderately disadvantaged communities showed increased uptake (aOR=1.70, 95% Crl=1.46-1.94, aOR=2.15, 95% Crl=1.89-3.08, and aOR=1.84, 95% Crl=1.63-2.13, respectively).

**Conclusions:** Community-level factors such as urban residence, female-headed households, and residence in the least disadvantaged communities were also linked to higher IPTp-SP uptake. Optimal uptake of IPTp-SP was more evident in the least disadvantaged regions. The findings of this study should guide the Cameroonian Ministry of Health and other stakeholders in developing targeted interventions to enhance the uptake of IPTp-SP in Cameroon.

## Background

Malaria during pregnancy remains a significant global health concern that can have severe consequences for both the mother and the developing fetus. However, cost-effective interventions have been available for more than two decades that aim to mitigate these harmful effects (Moore BR, Davis TME, 2018; Hill et al., 2013; Menéndez, D’Alessandro, ter Kuile, 2007). In 2004, the World Health Organization (WHO) recommended a comprehensive strategy for the prevention and management of malaria in pregnancy, which includes the administration of intermittent preventive treatment with sulfadoxine-pyrimethamine (IPTp-SP), the use of insecticide-treated nets (ITNs), and the effective management of clinical malaria and anemia (WHO, 2013).

Numerous investigations have demonstrated that malaria infection is highly endemic throughout Africa. The World Health Organization (WHO) reported the prevalence of malaria in Africa, revealing that approximately 80% of the 207 million clinical cases of malaria worldwide were recorded in African regions, and approximately 90% of the 627 thousand global malaria deaths were reported in these same regions in 2013. Within Africa, sub-Saharan Africa bears the greatest burden of malaria, with children under the age of five and pregnant women facing the highest risk of infection. Cameroon represents one of the 15 highest contributors to malaria incidence in sub-Saharan Africa and accounted for 3% of all global malaria cases in 2018, ranking as the third-highest incidence of malaria in Central Africa in 2019 (WHO, 2019). Cameroon has implemented a comprehensive approach to combat malaria that includes five complementary interventions, including indoor residual spraying, seasonal malaria chemoprevention, intermittent preventive treatment for pregnant women, insecticide-treated net distribution, and case management (Antonio-Nkondjio et al., 2019). Cameroon was also among the countries that received support from the US government’s President’s Malaria Initiative 2009-2014 to control and eliminate malaria (Antonio-Nkondjio et al., 2019).

Recent scientific research highlights the alarming reduction in uptake of intermittent preventive treatment during pregnancy (IPTp), leading to an increased risk of malaria among expectant mothers (Dun-Dery et al., 2021; Pons-Duran et al., 2021). To combat this issue, the World Health Organization (WHO) recommends the administration of IPTp using IPTp-SP in areas with moderate to high malaria transmission rates in Africa. WHO advises that pregnant women should receive at least three doses of SP, beginning as early as possible in the second trimester, with each dose administered at least one month apart and continued until delivery (WHO, 2013).

To address the low uptake of IPTp, WHO has released new guidelines on antenatal care in 2021, emphasizing the importance of increasing the number of contacts between healthcare providers and pregnant women (WHO, 2021). These updated guidelines offer a promising solution to expand the coverage of IPTp-SP by providing more opportunities for pregnant women to receive preventive treatment. An inter-agency briefing note published alongside the guidelines includes an ANC contact schedule with proposed timelines for the implementation of IPTp, which can be adapted to meet the unique needs of each country.

According to routine data from the malaria information system, 81% of pregnant women attending antenatal care (ANC) in Cameroon received at least one dose, 62% received at least two doses and 40% received at least three doses of IPTp after implementation of a IPTp-SP policy that urged expectant mothers to take-up least three free SP doses between the 16th and the 36th weeks of pregnancy (President’s Malaria Initiative Cameroon, 2018; Anchang-Kimbi et al 2014). In addition, the country has also implemented mass-campaign and free distribution of ITNs coupled with routine ANC distribution of ITN to complement the SP intervention.

Limited studies have been conducted on the on the uptake of IPTp in Cameroon since the inception of the policy limited. Most of the studies assessed the effectiveness of IPTp-SP interventions (Fokam, Ngimuh, Anchang-Kimbi, Wanji, 2016; Leonard, Eric, Judith, Samuel,2016; Hill, et al 2013). What is more worrying and necessitates this study is that regardless of the investment into addressing malaria cases, malaria cases remain one of the main causes of morbidity (52%) and mortality (14%) particularly in children under five years in Cameroon (WHO, 2013). This study sought to assess the factors that determine IPTp-SP uptake in Cameroon.

## Methods

### Data source

The study data utilized in this research was obtained from the 2018 CDHS, which used a two-stage sampling design to systematically select clusters or enumeration areas (EAs) and households for the survey. The Cameroonian National Institute of Statistics (NIS) and the United States Agency for International Development (USAID) collaborated with other international and local institutions to conduct the 2018 Cameroon Demographic and Health Survey (CDHS), which is part of the cross-country Demographic and Health Survey (DHS) project that covers approximately 85 low- and middle-income countries worldwide. In the first stage, the probability proportional sampling technique was employed to determine the sample size of EAs, while the second stage involved the selection of households. The survey successfully interviewed 13,527 women in the 15-49 age cohort, and the methodological procedure is detailed in the final report of the survey. This study focused on a subset of 4,823 eligible women from the survey dataset.

### Derivation of variables

#### Outcome variable

Optimal uptake of IPTp-SP was the outcome variable. In the 2018 CDHS, all women with birth history were asked if they had IPTp-SP, also termed as Fansidar, and to state the times they took it during the last pregnancy. According to the WHO, all women in moderate to high malaria transmission areas including Africa are to have a minimum of three doses of IPTp-SP (WHO, 2013 WHO; 2010). Following this recommendation, research participants who had three or more doses of IPTp-SP were classified to have had optimum IPTp-SP (labelled as 1). Women with less than three doses were classified as not having optimum IPTp-SP (labelled as 0).

#### Explanatory variables

To determine the optimal uptake of IPTp-SP, we considered thirteen explanatory variables. In line with previous DHS studies, we classified these variables into three levels: individual, community, and region (Yaya, Uthman, Amouzou, Bishwajit. 2018; Larsen, Merlo,2005). At the individual level, we examined age (15–19, 20–24, 25–29, 30–34. 35–39, 40–44, 45-49), educational attainment (no education, primary, secondary, higher), wealth quintile (poor, poorer, middle, richer richest), marital status (single, married, cohabiting, divorced, widowed), birth order (1, 2, 3), number of antenatal care visits (0, less than 8 (<8), 8 or more (≥8)), desire for last child (wanted then, wanted later, wanted no more), possession of insecticide treated net (no, yes), and possession of health insurance (no, yes). At the community level, we analyzed residents (urban, rural), socio-economic disadvantage (tertile 1 [least disadvantaged], 2 [moderately disadvantaged], and tertile 3 [most disadvantaged]), and household head (male, female).

Finally, at the regional level, we examined socio-economic disadvantage. Socio-economic disadvantage was determined based on the proportion of illiterate and poor individuals within the sample. The hierarchical nature of our study allowed us to effectively assess the impact of these factors on IPTp-SP uptake.

#### Analytical procedure

In this study, we utilized Stata version 13 to perform various analyses. The first step was to conduct a descriptive computation of the frequency and percentage of women who reported optimum IPTp-SP uptake during their last pregnancies. To investigate the association between explanatory variables and optimum IPTp-SP uptake, we employed the chi-square test at a 95% significance level. Table 1 presents the outcome of all the descriptive analyses. Next, we utilized a multi-level binary logistic regression with four models to inferentially analyze the data. Model I, the empty model, decomposed the variance existing between the community and regional levels. Model II comprised individual-level factors, whereas Model III reflected individual and community-level factors. The final and complete model, Model IV, reflected individual, community, and region-level factors. We calculated the adjusted odds ratios (AaOR) of the fixed effects with a 95% credible interval (CrI). Furthermore, we gauged the random effects by estimating the Intraclass Correlation (ICC) and median odds ratio (MOR). The ICC indicates the percentage of the overall variance in the probability of optimum IPTp-SP uptake that is associated with the clustering of odds of optimum IPTp-SP uptake within the same community or region. The MOR estimates the community or regional level variance as odds ratio and calculates the probability of optimum IPTp-SP uptake that is attributable to community and regional contexts (Yaya, Uthman, Amouzou, Bishwajit. 2018).

**Table 1:** Distribution of sample by IPTp-SP utilization in last pregnancy.

| Variable | At least 3 doses of IPTp-SP in last Pregnancy |  |  | X<sup>2</sup>; p-value |
| --- | --- | --- | --- | --- |
|  | No | Yes | Total |  |
|  | n (%) | n (%) | n (%) |  |
|  | 2886(59.8) | 1937(40.2) | 4823(100) |  |
| **Individual level** |  |  |  |  |
| Age |  |  |  | 77.562; p<0.05 |
| 15-19 | 273(61.1) | 174(38.9) | 447(100) |  |
| 20-24 | 673(62.4) | 405(37.6) | 1078(100) |  |
| 25-29 | 758(58.7) | 533(41.3) | 1291(100) |  |
| 30-34 | 617(60.2) | 407(39.8) | 1024(100) |  |
| 35-39 | 365(56.6) | 279(43.4) | 644(100) |  |
| 40-44 | 168(58.6) | 119(41.4) | 287(100) |  |
| 45-49 | 31(61.9) | 19(39.1) | 51(100) |  |
| Education |  |  |  | 24.263; p<0.001 |
| No education | 508(56.1) | 398(43.9) | 905(100) |  |
| Primary | 932(63.9) | 526(36.1) | 1458(100) |  |
| Secondary | 1268(60.0) | 848(40.1) | 2116(100) |  |
| Higher | 178(51.9) | 165(48.1) | 344(100) |  |
| Wealth quintile |  |  |  | 43.375; p<0.001 |
| Poorest | 391(55.8) | 310(44.2) | 701(100) |  |
| Poorer | 644(65.3) | 342(34.7) | 986(100) |  |
| Middle | 668(65.2) | 356(34.8) | 1024(100) |  |
| Richer | 645(58.0) | 467(42.0) | 1112(100) |  |
| Richest | 539(53.8) | 462(46.0) | 1001(100) |  |
| Marital status |  |  |  |  |
| Single | 523(58.8) | 366(41.1) | 889(100) | 23.375; p<0.05 |
| Married | 1665(58.8) | 1167(41.2) | 2832(100) |  |
| Cohabiting | 624(64.1) | 349(35.9) | 973(100) |  |
| Divorced | 34(69.1) | 15(30.9) | 49(100) |  |
| Widowed | 39(50.1) | 39(49.9) | 78(100) |  |
| Birth order |  |  |  |  |
| 1 | 1184(58.3) | 847(41.7) | 2030(100) | 1.443; 0.486 |
| 2 | 1166(60.9) | 748(39.1) | 1914(100) |  |
| 3 | 536(61.1) | 341(38.9) | 877(100) |  |
| ANC visit |  |  |  | 18.428; p<0.001 |
| 0 | 85(68.9) | 38(31.1) |  |  |
| <8 | 2572(60.3) | 1694(39.7) |  |  |
| ≥8 | 229(52.8) | 204(47.2) |  |  |
| Wanted last child |  |  |  | 22.428; p<0.01 |
| Wanted then | 2177(59.2) | 1498(40.8) | 3675(100) |  |
| Wanted later | 584(62.5) | 350(37.5) | 934(100) |  |
| Wanted no more | 125(58.4) | 89(41.6) | 214(100) |  |
| Have ITN |  |  |  | 6.109; p<0.05 |
| No | 526(64.3) | 2911(35.7) | 817(100) |  |
| Yes | 2360(58.9) | 1646(41.1) | 4005(100) |  |
| Health insurance |  |  |  | 5.587; p<0.05 |
| No | 2837(60.1) | 1887(39.9) | 4724(100) |  |
| Yes | 49(49.4) | 50(50.6) | 99(100) |  |
| **Community level factors** |  |  |  |  |
| Residence |  |  |  | 28.295; p<0.001 |
| Urban | 1476(57.2) | 1103(42.8) | 2579(100) |  |
| Rural | 1410(62.8) | 834(37.2) | 2244(100) |  |
| Socio-economic disadvantage |  |  |  | 20.205; p<0.01 |
| Tertile 1(least disadvantaged) | 857(59.2) | 590(40.8) | 1447(100) |  |
| **Tertile 2** | 1001(62.5) | 560(37.5) | 1600(100) |  |
| **Tertile 3(most disadvantaged)** | 1028(57.9) | 748(42.1) | 1776(100) |  |
| Household head |  |  |  | 3.968; p<0.05 |
| Male | 2296(60.9) | 1471(39.1) | 3767(100) |  |
| Female | 590(55.9) | 466(44.1) | 1056(100) |  |
| **Region level factors** |  |  |  |  |
| Socio-economic disadvantage |  |  |  | 14.086; p<0.01 |
| Tertile 1(least disadvantaged) | 924(58.7) | 649(41.3) | 1573(100) |  |
| Tertile 2 | 1175(64.4) | 649(35.6) | 1824(100) |  |
| Tertile 3(most disadvantaged) | 787(55.2) | 639(44.8) | 1426(100) |  |

#### Model fit and specifications

To ensure the robustness and reliability of our estimations, we implemented several precautionary measures. We utilized the Variance Inflation Factor (VIF) technique, as proposed by Akinwande, Dikko, and Samson (2017), to examine the presence of multicollinearity among the explanatory variables. Our analysis indicated that there were no substantial correlations among the variables (mean VIF=1.49, minimum VIF=1.09, maximum VIF=2.48). To evaluate the appropriateness of our models, we employed the Bayesian Deviance Information Criterion (DIC) and Markov Chain Monte Carlo (MCMC) estimation. Finally, we fitted all models using the MLwinN package version 3.05 of the Stata software.

#### Ethics approval

Based on pre-existing data from the 2018 CDHS, this study was conducted with the approval of the ethics committee of ORC Macro Inc. The procedures were performed in accordance with the regulations for the protection of human subjects outlined by the U.S. Department of Health and Human Services, as verified by the Inner-City Fund (ICF) International. The full ethical protocols can be accessed at http://goo.gl/ny8T6X. As secondary data were used, informed consent was not required, and all consent protocols were previously addressed by the institutions that collected the data. Further information regarding the protection of respondents’ privacy during the DHS survey can be found at https://dhsprogram.com/methodology/Protecting-the-Privacy-of-DHS-Survey-Respondents.cfm.

#### Patient and Public Involvement

No patient involved.

### Results

#### Descriptive findings

Results 40.2% of the surveyed women had at least three doses of IPTp-SP and attainment of three or more doses was phenomenal among women aged 35-39 (43.4%) and those having higher education (48.1%) (Table 1). Forty-six percent (46%) of women in the richest quintile women had at least three doses whilst nearly half of widowed women did the same during their last pregnancies (49.9%). A significant proportion of women at first birth order had optimum IPTp-SP (41.7%) just as women who obtained more than eight (8) ANC visits during their last pregnancies (47.2%). Attainment of three IPTp-SP doses or higher was phenomenal among women who wanted no more children (41.6%) and those with ITN (41.1%). Half of those with health insurance had optimum IPTp-SP (50.6%). Optimum IPTp-SP uptake was also substantial among urban residents (42.8%), most disadvantaged women at the community (42.1%) and the region level (44.8%), as well as women with female household heads (44.1%).

Table 2 presents the results of the random effects analysis. The final model (Model IV) indicates that women aged 35-39 had higher odds of receiving at least 3 doses of IPTp-SP during their last pregnancies compared to those aged 45-49 (adjusted odds ratio [aOR]=1.92, 95% credible interval [Crl]=1.08-3.41). However, women with no formal education had lower odds of attaining optimal IPTp-SP uptake (aOR=0.81, 95% Crl=0.70-0.92) compared to those with higher education. Additionally, women who were categorized as the richest (aOR=1.47, 95% Crl=1.20-2.09) and those who attended at least eight antenatal care (ANC) visits (aOR=1.97, 95% Crl=1.25-3.12) had higher odds of attaining optimal IPTp-SP uptake than those categorized as the poorest and those who attended less than eight ANC visits, respectively. Women with health insurance (aOR=1.70, 95% Crl=1.46-1.94), urban residence (aOR=2.15, 95% Crl=1.89-3.08), and residents of moderately disadvantaged communities (aOR=1.84, 95% Crl=1.63-2.13) had higher odds of receiving three or more IPTp-SP doses compared to women without health insurance, rural residents, and residents of the most disadvantaged communities. Furthermore, our findings suggest that women whose household heads were male had lower odds of achieving optimal IPTp-SP uptake (aOR=0.84, 95% Crl=0.72-0.99), while women residing in at least disadvantaged regions had higher odds of receiving at least three IPTp-SP doses compared to those residing in the most disadvantaged regions (aOR=1.69, 95% Crl=1.44-2.06).

**Table 2:**
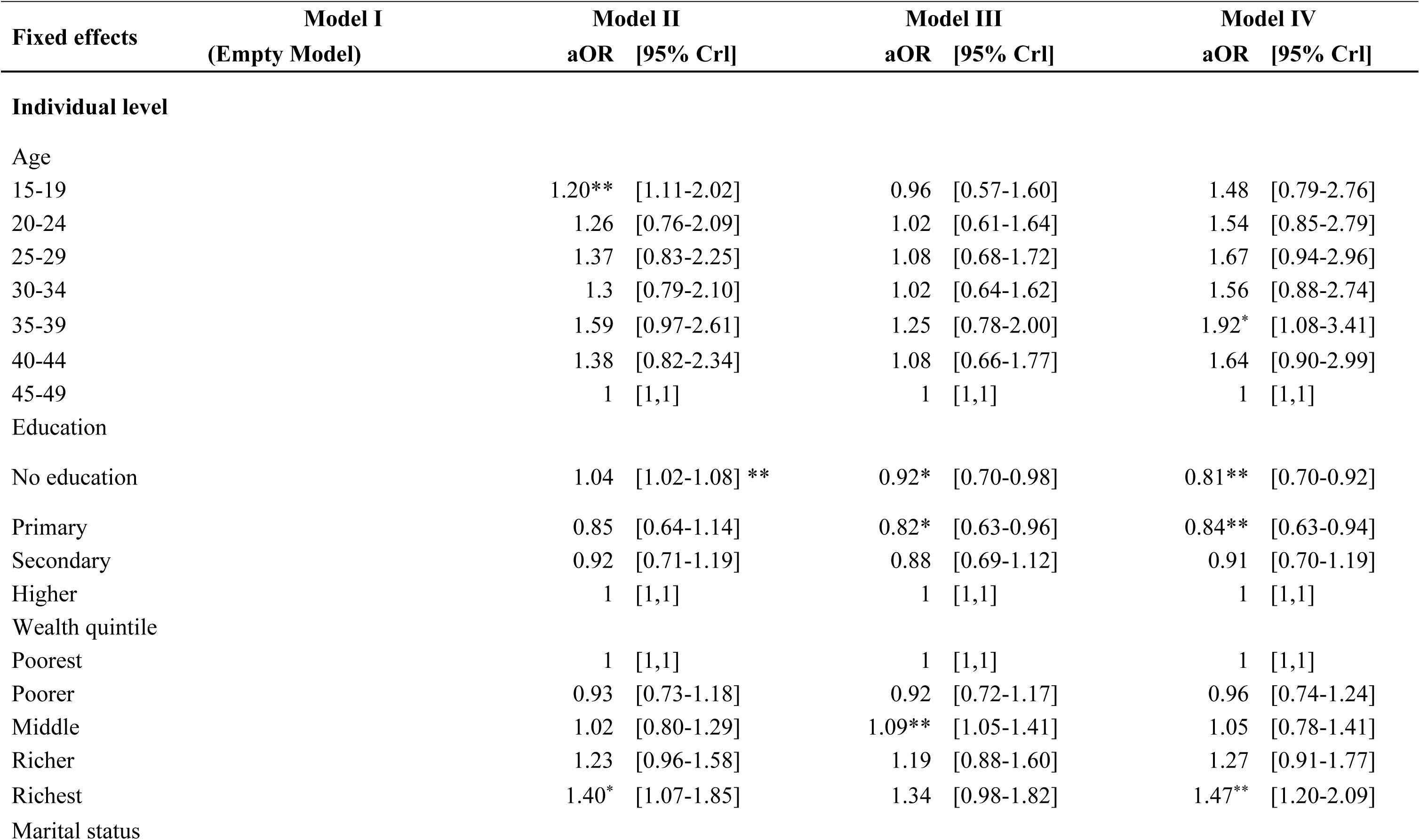
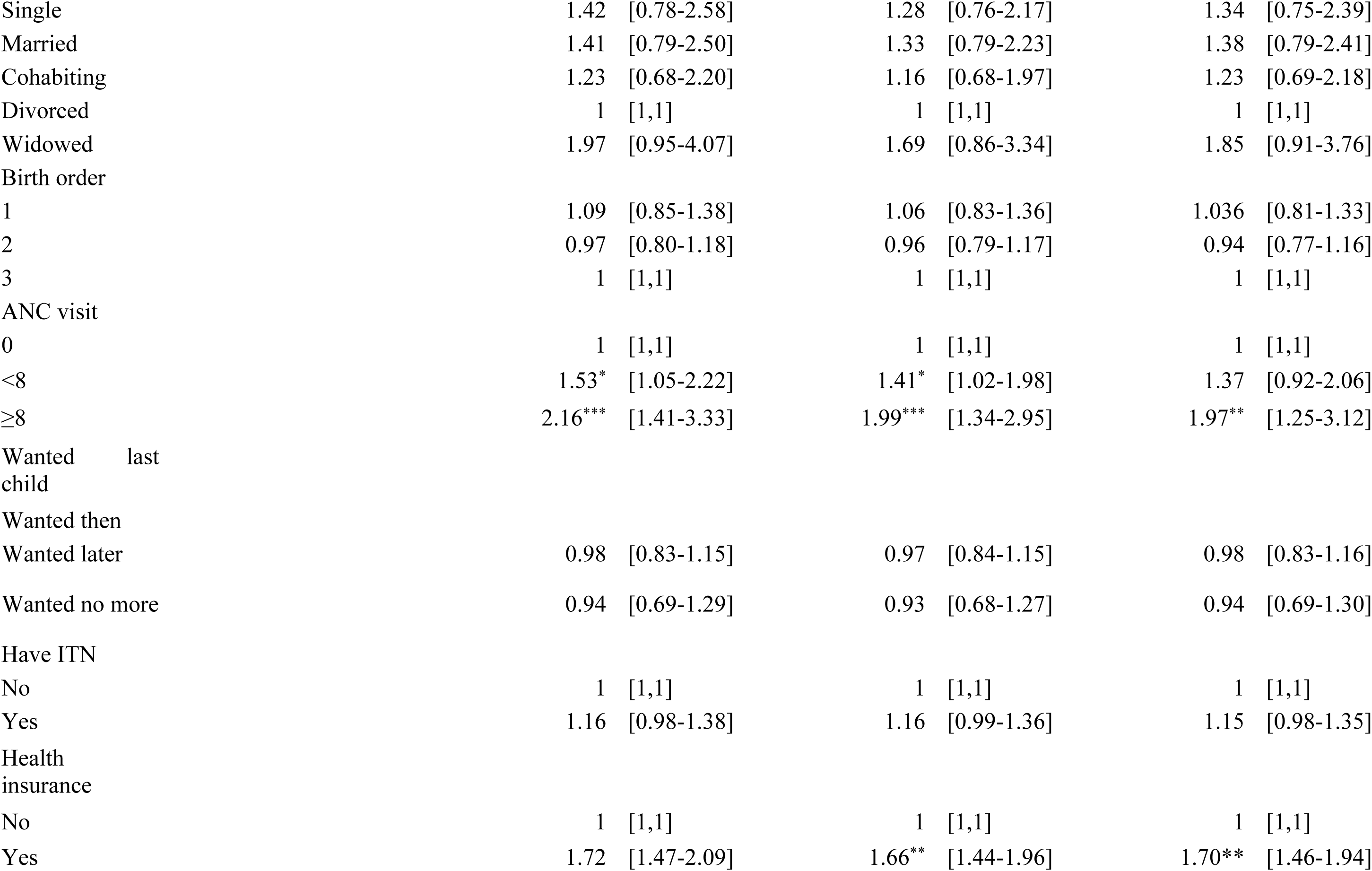
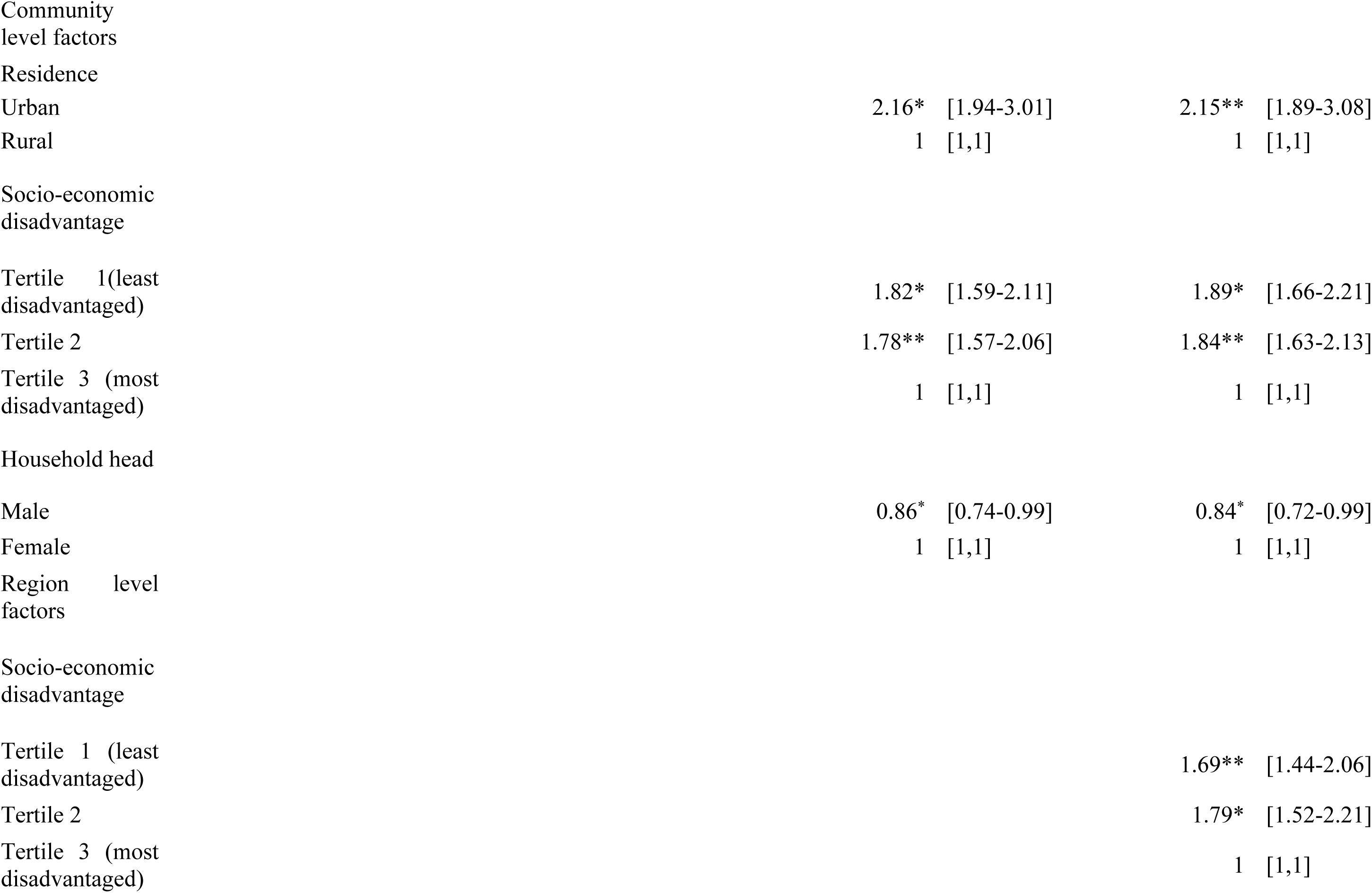
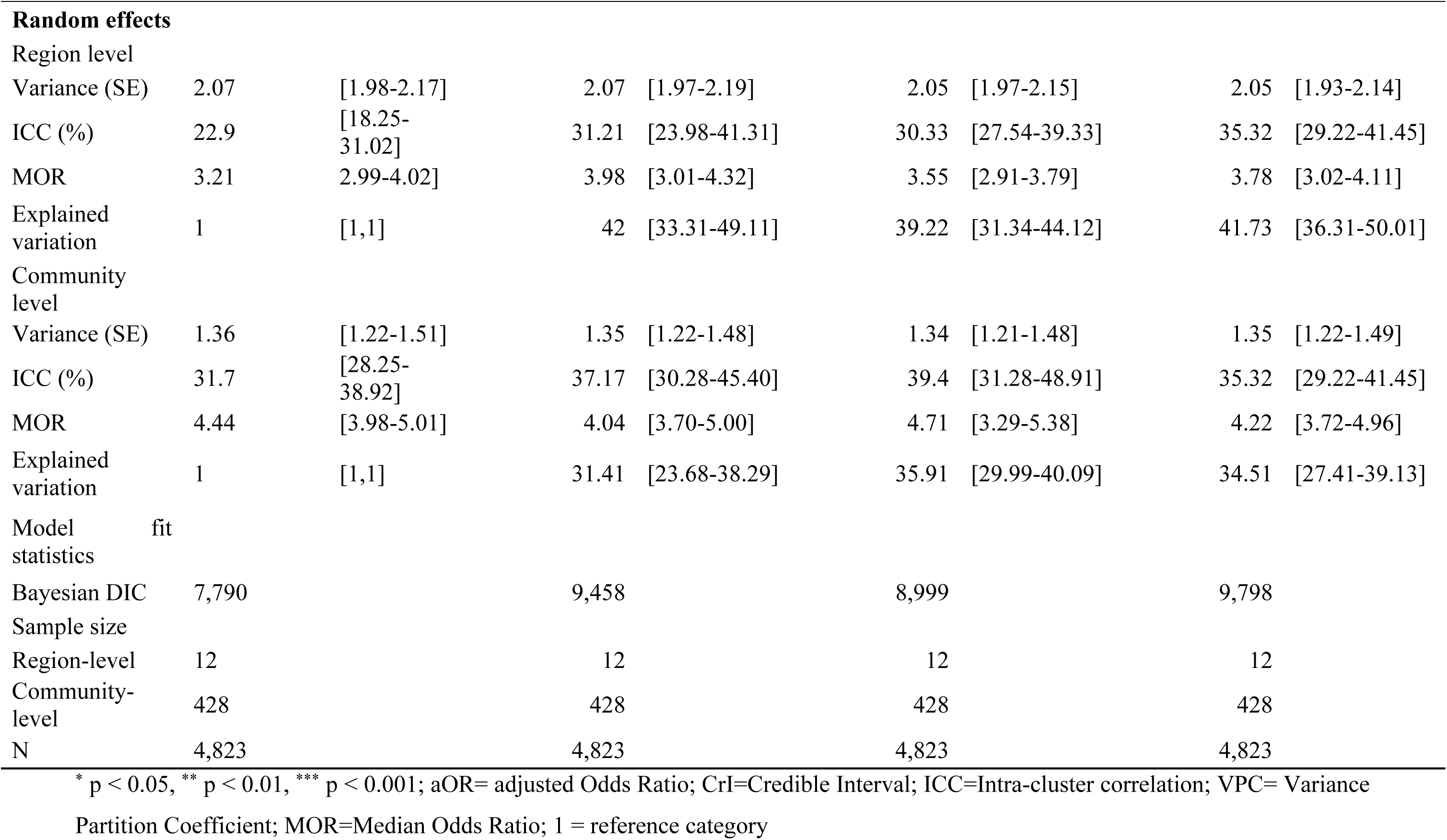
Table 2. Individual, community and region-level factors associated with optimum IPTp-SP uptake.

#### Random effects’ results

The present study utilized a random effects model. The finding from table 2 shows that significant variation exits across the 12 regions [σ2 = 2.07, 95% Crl=1.98-2.17] and the 428 communities (σ2 = 1.36, Crl=1.22-1.51). The model’s ICC estimates of 22.90% (for region) and 31.70% (for communities) suggest that factors at the region and community levels significantly impact the likelihood of optimum IPTp-SP uptake. The results from the MOR analysis indicate that both region and community factors influence the uptake of IPTp-SP. The final model indicates that when a woman relocates from one region to another, her likelihood of optimal IPTp-SP uptake increases by a median of 3.78% [3.02-4.11]. Moreover, women are more likely to have optimum IPTp-SP uptake when they move from a community with lower probability of uptake to one with higher probability of uptake, with a median increase of 4.22% [3.72-4.96] in their odds of uptake.

## Discussion

The focus of this study was to assess the factors that determine IPTp-Sp uptake in Cameroon. From our study, the uptake of optimal IPTp-SP was found to be positively related to one’s level of education. The higher one’s level of education the higher the chances of attaining the optimal level of IPTp-SP uptake. This finding corroborates Exavery et al. (2014) and the report of WHO, (2019) that found an increase in the chances of optimal uptake of IPTp-SP the higher one rises in their level of education. The results imply that supporting women’s education beyond primary education serves as a major catalyst to decision making autonomy against restrictive traditions. It also affirms the view that education empowers women in their healthcare decision making process.

It was also revealed that pregnant women who attended at least eight ANC visits are more likely to attain optimal doses IPTp-SP. Although not within the scope of the paper to find the reasons for this observation, the more contact a pregnant woman has with the health staff appeared to increase chances of attaining the optimal doses of IPTp-SP. This corroborates Amakwah and Anto, (2019), Oppong et al. (2019), Bouyou-Akotet et al. (2013), and Anchang-Kimbi et al. (2014) studies. These studies noted that early initiation of ANC visits creates the opportunity for the pregnant woman to have more contact time with midwives (and the caregivers) who comprehensively assessed and provided all the package of services including IPTp-SP. A couple of factors play a major role in determining the number of visits a pregnant woman makes including her socioeconomic status (Anchang-Kimbi et al 2014), employment status, and husband’s educational status (Sakeah, 2017). Exavery et al. (2014) had a contrary observation. They argued that quality antenatal services are more important than the count of visits that do not guarantee quality services. One’s health insurance status emerged as one of the key determinants of uptake of IPTp-SP by pregnant women in the present study. However, previous studies have not reported the relationship between having health insurance and optimal uptake of IPTp-SP. Although IPTp-SP services in Cameroon is free at the service delivery point (Dionne-Odom, 2017), other cost associated with ANC services could hinder women’s visit to the health facility to attend ANC services. Pregnant women with health insurance access these services through the insurance, thereby having an increased likely to seeking antenatal care and benefit from SP services.

According to the study, urban women and those from moderately disadvantaged communities had higher chances of receiving three or more doses of IPTp-SP during pregnancy than rural women who usually resided in the most disadvantaged communities. Furthermore, women attending rural health facilities were less likely to complete the recommended SP doses during pregnancy. This finding is consistent with previous studies by Azizi et al. (2019) and Onoka et al. (2019), which found that women receiving IPT-p/SP from rural health facilities in Nigeria were less likely to receive at least three doses of SP than urban women. Additionally, Martin et al. (2013) observed that attending ANC at a lower-level health center relative to a hospital was associated with reduced likelihood of receiving optimal doses of IPTp-SP. Conversely, Arnaldo et al. (2017) reported that the overall prevalence of maternal malaria (peripheral and/or placental) was lower and higher among women from rural areas compared to those from urban areas. Some scholars have linked the effect of one’s residence to the rate of taking SPs to the fact that rural health facilities had lower stock levels of SP than urban health facilities due to poor supply chain management between district health offices and rural areas, which was caused by challenges in transporting the commodity or understaffing (Dionne-Odom, Westfall, Apinjoh, Anchang-Kimbi, Achidi, Tita, 2017; Hill et al, 2013). This resulted in high client-to-staff ratios and subsequently long queues and waiting times.

Several maternal sociodemographic characteristics were found to be associated with the uptake of at least one dose of IPTp-SP. With regards to household heads and IPTp uptake, we noted that women in male headed households were less likely to achieve optimal IPTp-SP compared to women in female headed households. This may be due to the low knowledge and education these household heads have on the benefits of IPTp-SP. Again, men who have their wives sharing or having had experience of drug reactions will also be reluctant or turn to influence their wives from either starting or completing the uptake of IPTp dose. This thought follows Tobin-West and Asuquo (2018) who posit that women who share misconceptions about chloroquine dispensing in the past, despite a change in treatment policy by recommending the use of SP instead of chloroquine, are challenged by their husband in their (women’s) decision to utilize IPTp-SP when they attend ANC together. Molyneux et al. (2012) also assert that the unawareness level of some male headed households about the importance of the medicine is more likely to influence men’s decision to stop their wives from using the medicine. from either starting or completing the uptake of IPTp dose.

According to scientific research, residents of the least disadvantaged regions are more likely to receive at least three doses of IPTp-SP, in contrast to those in the most disadvantaged regions. This indicates that countries with robust economies, where the majority of people belong to the wealthiest wealth quintile, are likely to have high IPTp-SP uptake rates. Yaya et al. (2020)31 support this argument by noting that women residing in poorer households had a significantly higher chance of not receiving at least three doses of IPTp-SP during their last pregnancy, compared to those residing in the richest households. However, this finding does not imply that low-income regions in Cameroon and other sub-Saharan African (SSA) countries are unable to combat malaria during pregnancy using IPTp-SP. Instead, it highlights the importance of prioritizing and increasing investments in malaria in pregnancy (MiP) interventions in low-income regions of Cameroon to ensure the well-being of pregnant women and their newborns.

## Conclusion and policy implications

Low uptake of optimal IPTp-SP remains a significant public health challenge in Cameroon, necessitating a concerted effort to improve the situation. This study aimed to identify the determinants of IPTp-SP uptake in Cameroon. Results at the individual level suggest that women’s educational status, number of antenatal care visits, possession of health insurance, and being in the highest wealth quintile were positively associated with optimal IPTp-SP uptake. Community-level factors such as urban residence, female-headed households, and residence in the least disadvantaged communities were also linked to higher IPTp-SP uptake. Optimal uptake of IPTp-SP was more evident in the least disadvantaged regions. The findings of this study should guide the Cameroonian Ministry of Health and other stakeholders in developing targeted interventions to enhance the uptake of IPTp-SP in Cameroon.

## Data Availability

The datasets are publicly available in the DHS website https://dhsprogram.com/

## Author contributions

LB conceived the study, performed the analysis and developed the methods. AN and MHK drafted the background. AMT and JPKN worked on the discussion. KAA and MMZ, reviewed multiple drafts, and proposed additions and changes. All authors have reviewed and approved the final version of the manuscript.

## Acknowledgements

We appreciate the Measure DHS for granting us data for this study.

